# A blinded, controlled randomized clinical trial on the efficacy of neck muscle vibration in patients with post-stroke neglect

**DOI:** 10.1101/2025.06.30.25330533

**Authors:** Britta Stammler, Carina Thiel, Anne Lieb, Heike Meißner, Hans-Otto Karnath

**Author notes:** Correspondence should be addressed to: Hans-Otto Karnath, Center of Neurology, University of Tübingen, Hoppe-Seyler-Str. 3, D-72076 Tübingen Germany.

## Abstract

**Background and Aim:** Unilateral spatial neglect (UN) impairs patients’ ability to detect and respond to stimuli on the contralesional side, severely limiting functional recovery after right-hemispheric stroke. Neck muscle vibration (NMV) has been shown as a bottom-up, proprioceptive intervention to modulate spatial neglect. Although preliminary studies found promising effects, the isolated efficacy of NMV for neglect rehabilitation has not yet been tested in a randomized, blinded controlled trial. This study aimed to evaluate whether NMV alone improves neglect symptoms and activities of daily living.

**Methods:** Twenty patients with right-hemispheric stroke and UN were randomly assigned to receive either active or placebo NMV over two weeks (5 sessions/week). Both groups received 20-minute daily vibration sessions. Standard neglect therapy was withheld in the active group but administered in the placebo group. Assessments included standard neglect diagnostics (e.g., Letter Cancellation, Bells Test), the Free Exploration Test (FET), and two ADL-based measures (NET, CBS), conducted before, immediately after, and (for the NMV group) one month post-treatment.

**Results:** The active NMV group showed significant improvements in three of four standard neglect tests, exploration behavior (FET), and ADL performance, with effects remaining stable at one-month follow-up. The placebo group showed comparable gains in ADL outcomes but improved in one standard neglect test only. Between-group analyses revealed no statistically significant differences, suggesting similar efficacy of both interventions.

**Conclusion:** NMV alone yields clinically meaningful and lasting improvements in neglect symptoms and daily functioning, comparable to standard active exploration therapy. Its passive nature makes it a promising tool, especially for early rehabilitation.

## Introduction

Unilateral spatial neglect (UN) is a neurological disorder that typically occurs following right-hemispheric brain damage. It is estimated to affect approximately 30% of stroke patients ^1^. The underlying lesions usually involve regions of the perisylvian network, which plays a crucial role in spatial orientation in the right hemisphere ^2^. Damage to this network in patients with UN manifests as a strong continuing rightward orientation combined with neglect of the left side. These symptoms are associated with more severe motor deficits and delayed rehabilitation ^3^, ultimately reducing patients’ ability to perform activities of daily living and negatively impacting their quality of life in the long term ^4,5^.

There are three main therapeutic approaches for treating UN: top-down interventions, which focus on cognitive control of exploratory behavior; neuromodulatory techniques, which aim to inhibit or activate specific cortical networks; and bottom-up interventions, which use sensory stimulation to modulate altered spatial representation ^6–8^. Bottom-up approaches have the advantage of being largely independent of any cognitive and attentional deficits associated with spatial neglect ^7^ and do not require the patients to have insight into their condition and therefore be able to consciously cooperate ^6,8^.

Neck muscle vibration (NMV) is a bottom-up intervention that was extensively studied in the 1990s and early 2000s ^9,10^. Research has shown that left-sided NMV can modulate spatial bias in both healthy individuals and patients with UN. The effects of NMV result from the stimulation of muscle spindle afferents, which were thought to induce a reorganization of egocentric spatial reference systems in the brain, typically in the direction contralateral to the site of vibration ^11–14^. Specifically, studies have demonstrated a significant expansion of the explored visual space toward the contralesional side following left-sided NMV ^15^ combined with a leftward shift in the perceived straight-ahead (SSA) orientation ^12^ This shift facilitates improved perception and exploration of the neglected space. These effects cannot be explained solely by a general increase in arousal, as other forms of sensory stimulation, such as electrical stimulation, do not produce comparable results ^16,17^.

Previous studies have evaluated NMV in repeated application scenarios ^14,18–20^. Johannsen et al. (2003) examined six patients with neglect and found that a 20-minute daily NMV intervention over two weeks led to long-lasting symptom improvements, detectable up to 1.4 years after treatment. Kamada et al. (2011) investigated 11 patients and reported that just five minutes of NMV before occupational therapy sessions over two weeks resulted in significant and stable improvements, persisting for up to two weeks post-treatment. The combination of neck muscle vibration and voluntary arm movements has also been tested successfully - but so far only in a single case (Ceyte et al., 2019). Schindler et al. (2002) conducted a crossover study with 20 neglect patients to compare active exploration training with simultaneous NMV against exploration training alone. Both interventions consisted of 15 sessions of 45 minutes each. The combined treatment led to significant and sustained improvements lasting up to two months, with visible benefits in activities of daily living.

Despite these promising findings, no randomized controlled trial (RCT) has yet been conducted to evaluate the isolated efficacy of NMV. Previous studies either lacked a control group or combined NMV with other therapeutic approaches to treat spatial neglect, making it difficult to determine the specific effects of NVM. This gap in evidence will be addressed by the present study, which, for the first time, investigates the standalone effectiveness of NMV on neglect symptoms in a randomized, controlled, and blinded study design.

## Material and Methods

The study was preregistered in the WHO-approved public trial registry “German Clinical Trials Register” (DRKS-ID: DRKS00032376).

### Participants

Twenty consecutively admitted patients with right-sided stroke and spatial neglect participated after the exclusion of five patients, one who was transferred to another hospital and four because of spontaneous remission after one week. Patients were recruited from two different rehabilitation facilities (Schmieder-Klinik, Stuttgart-Gerlingen, Germany [n = 17 patients], Neurological Rehabilitation Center Quellenhof, Sana Kliniken AG, Bad Wildbad, Germany [n = 3 patients]) and were randomly assigned to either a group who received active NMV or a group with placebo NMV (n=10 each). Demographic and clinical details are given in Table 1. Structural imaging was acquired by computed tomography as part of the clinical routine procedure carried out for all stroke patients in the acute phase at stroke-onset. Patients with tumors or patients in whom scans revealed no obvious lesions were not included. All participants gave their informed consent to participate in the study, which was conducted in accordance with the ethical standards of the 1964 Declaration of Helsinki.

**Table 1.**
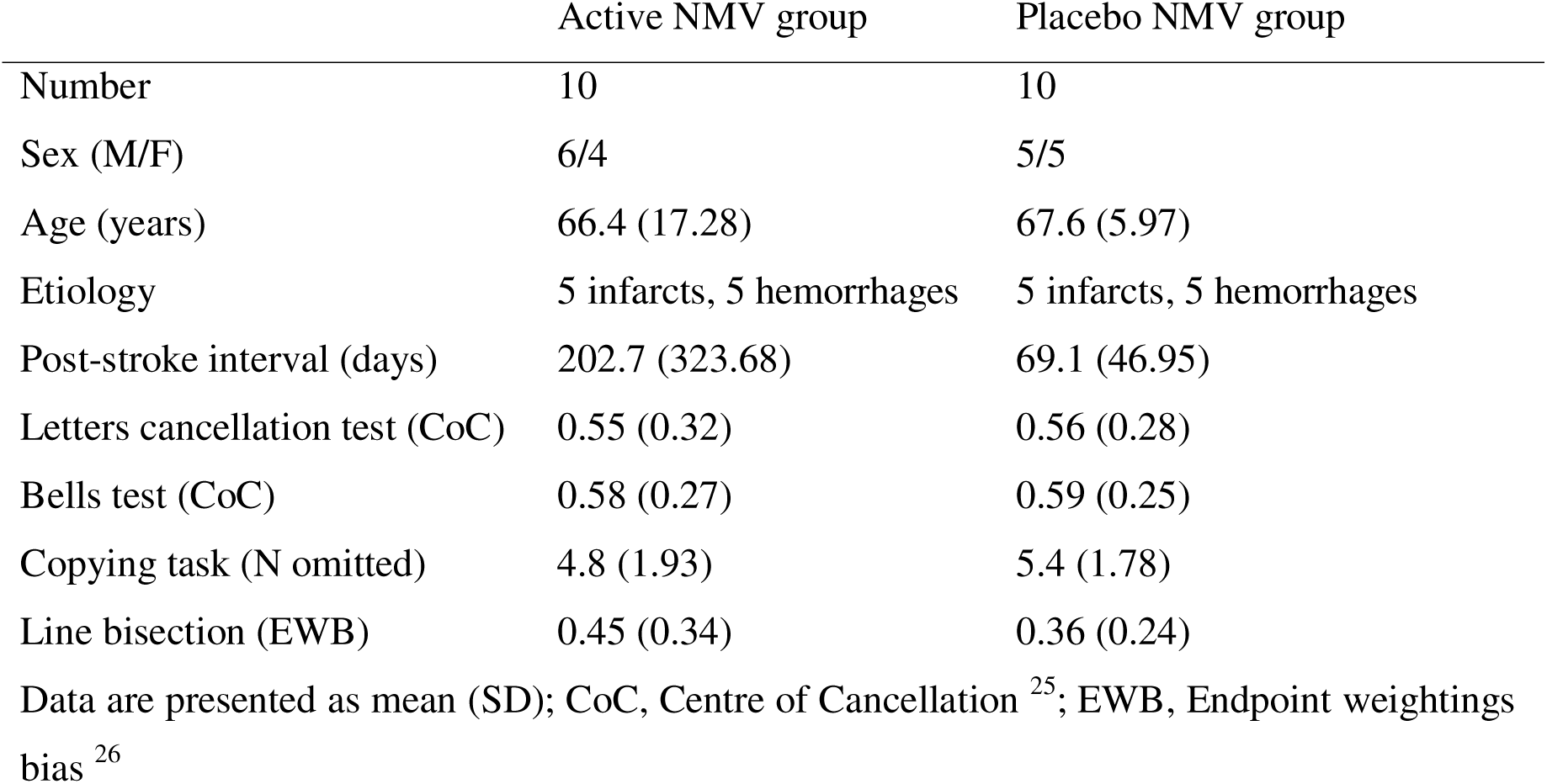
Demographic and clinical data of all 20 right brain damaged neglect patients.

The inclusion criterion for this study was the presence of spatial neglect as a result of brain damage caused by right hemispheric stroke. In addition to clinical behavioral observations, diagnostic criteria had to be met in at least two of the following four neglect tests: The Letter Cancellation Test ^21^, the Bells Test ^22^, a Copying Task ^23^, and a Line Bisection Task ^24^. All four neglect tests were performed on a Samsung S7+ tablet with screen dimensions 285x185mm. The severity of spatial neglect in the cancellation tasks was determined by calculating the center of gravity of the target stimuli marked in the search fields, i.e. the Center of Cancellation (CoC; ^25^. A CoC value ≥ 0.08 indicated left-sided spatial neglect ^25^. The Copying Task consisted of a complex scene consisting of four objects (fence, car, house, tree), points were assigned based on missing details or whole objects. One point was given for a missing detail, two for a whole object. The maximum number of points is therefore eight. A score higher than 1 (i.e. > 12.5% omissions) indicated spatial neglect ^23^. In the Line Bisection Task ^24^, patients were presented with four different line lengths eight times each, i.e. 32 lines in total. The cut-off value for spatial neglect was an ‘endpoint weightings bias (EWB)’ value ≥ 0.07 ^26^.

### Procedure

#### Intervention

Both groups, the active NMV group and the placebo NMV group, received daily 20-minute vibration sessions over a period of two weeks (five sessions per week). The vibration was applied using a handheld device (NOVAFON®, version: Novafon Power 2) equipped with a flat disk (diameter: 2.8 cm), which was placed on the left posterior neck muscles (Fig. 1A). The active NMV group received vibration at 100 Hz with an amplitude of 2.8 mm, whereas the placebo NMV group received an ineffective vibration with an amplitude of <0.09 mm, applied at the same neck region (Fig. 1A). To ensure blinding, the placebo device was identical in appearance to the active device. Instead of altering the frequency, the vibration was rendered ineffective by reducing the amplitude. It maintained a comparable humming sound and provided mild sensory stimulation, ensuring that participants perceived a similar experience in both groups. The participating patients were not informed about which group (active or placebo NMV) they were assigned to.

**Figure 1.**
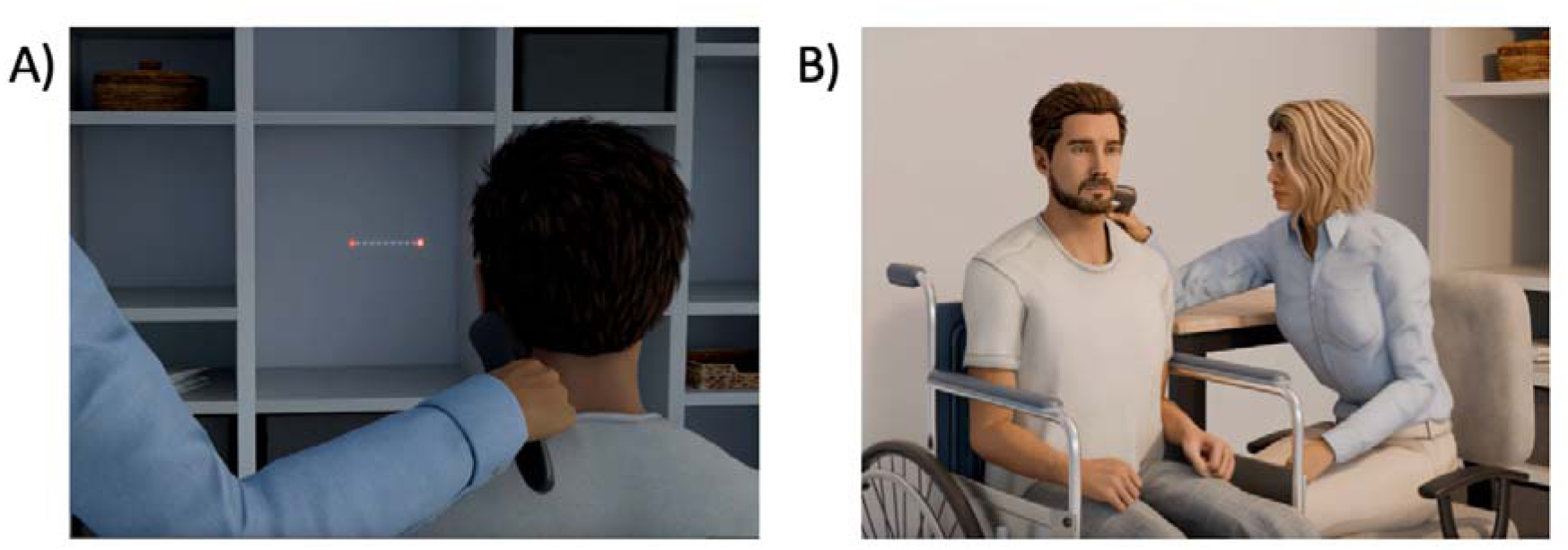
(A) Determination of the correct vibrator position in the active NMV group. The optimal position was identified as the location where the patient experienced - in dimmed lighting - the strongest illusion of a horizontal displacement of a centrally presented stationary red light towards the right side. (B) Setup for vibration of left posterior neck muscles in both the active NMV group as well the placebo NMV group during the treatment sessions.

In all patients, the vibrator was positioned within a palpable depression in the left dorsolateral neck region. Based on anatomical orientation, this site corresponds to the upper part of the so-called ‘posterior triangle of the neck’, i.e. the area of the depression between the left sternocleidomastoid muscle and the upper, descending part of the left trapezius muscle. In the active NMV group, exact positioning of the vibration device within that region was further individually adjusted to maximize the vibration-induced shift in the perceived body midline. To determine the optimal placement, adjustments were made in dimmed lighting until the subject experienced the strongest illusion of a rightward horizontal displacement of a centrally presented stationary red light (cf. Fig. 1A). Of the ten patients in the active NMV group, five reported experiencing such a kinesthetic illusion of the centrally presented light stimulus. Once the ideal location was identified, it was marked with a permanent marker to ensure consistent placement during subsequent treatment sessions, which were conducted under normal lighting conditions (cf. Fig. 1B).

In addition, both groups (active and placebo NMV) received standard computer-based cognitive training of the respective rehabilitation institution, as part of the neuropsychological rehabilitation program. This consisted in both facilities of five training sessions per week, each session lasted about 25 minutes. The experimental NMV group received this training without any neglect-specific modules, while this training in the placebo NMV group included either neglect-specific computer training modules or individual therapy sessions focusing on smooth pursuit eye movement training and visual exploration therapy. For smooth pursuit eye movement training, control patients were presented with clouds of dots on a computer screen that moved slowly at 5-10°/s from the ipsilesional to the contralesional side. Patients were instructed to follow the moving dots with their eyes. For visual exploration therapy, patients completed reading and copying tasks, image descriptions, and search tasks, all of which required them to actively orient toward their contralesional side.

### Assessment of treatment effects

A comparison of the active NMV group against the placebo NMV group allowed to directly compare the effect of pure NMV therapy against the effect of standard neglect therapy. Before and after the intervention, both groups underwent a total of four examinations (cf. Fig. 2) using the four diagnostic neglect tests described above (Letter Cancellation, Bells Test, Copying Task, Line Bisection Task), as well as the Free Exploration Test (FET; ^27^, and everyday tasks from the German version of the Behavioral Inattention Test (BIT, German version: Neglect Test (NET; ^28^) and the Catherine Bergego Scale (CBS; ^29^. Two of the five diagnostic assessments were conducted before the start of the intervention to account for spontaneous recovery (Fig. 2; E1 and E2). After completing the intervention, both patient groups underwent two additional assessments: immediately after the training and one week post-training (Fig. 2; E3 and E4). Additionally, the active NMV group received a fifth follow-up assessment one month after completing the training (Fig. 1; E5).

**Figure 2.**
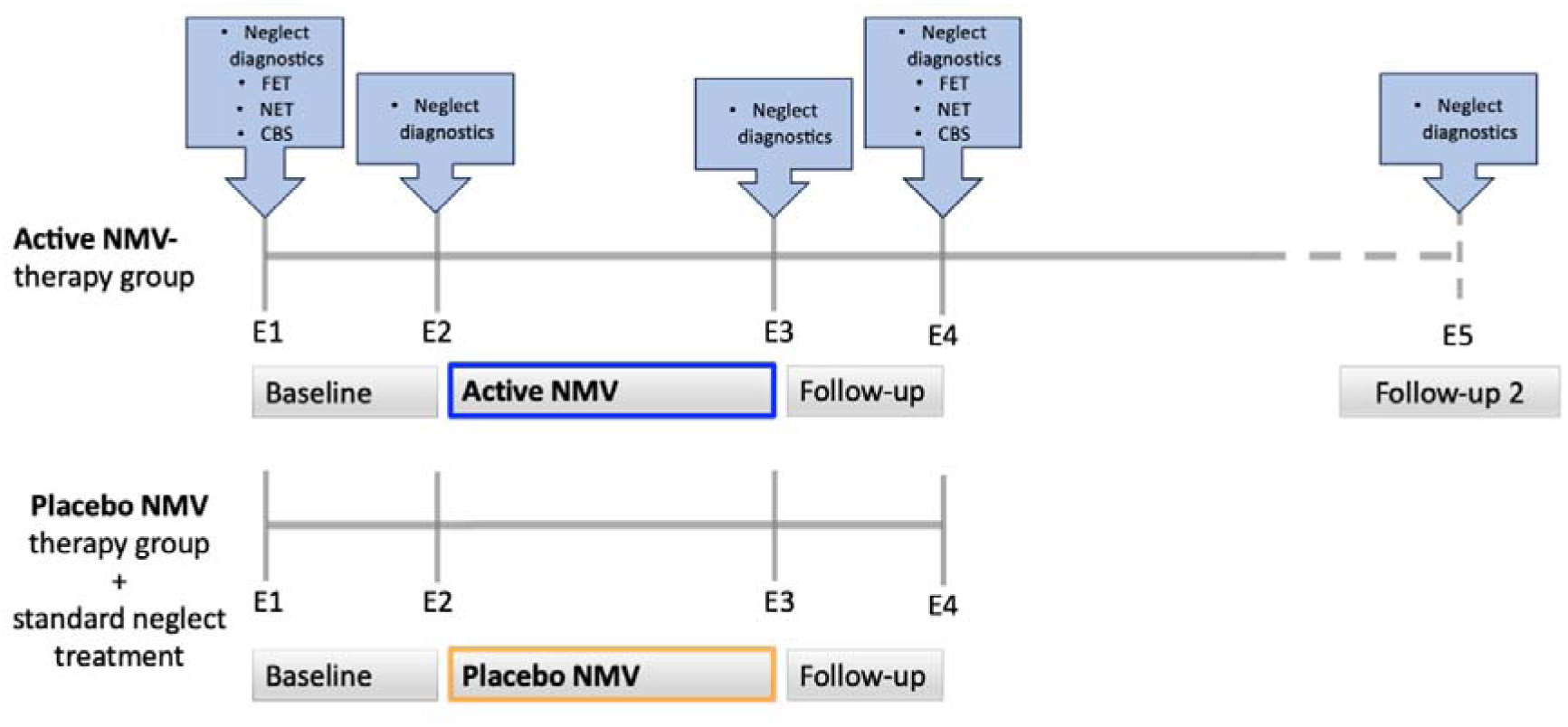
Experimental design. Four diagnostic examinations (E1 to E4) were performed in both the active NMV group and the placebo NVM group. Additionally, the active NMV group was examined one month after completion of therapy. The four diagnostic neglect tests were performed at all examination time. At time points E1 and E4, additionally the FET and the two ADL tests NET and CBS were carried out.

To assess activities of daily living (ADL), 8 out of the 17 tasks from the BIT/NET were selected. These tasks included reading a newspaper article, writing an address, describing a picture of a meal, a sink, and a room, reading a menu, and reading both a digital and an analog clock. As a second method for assessing ADL, the Catherine Bergego Scale (CBS) was conducted as an observer-rated assessment. The CBS evaluates functional impairments in personal care, spatial exploration, and navigation, based on observations made by caregivers or healthcare professionals. It consists of 10 items, each rated on a 4-point scale (0–3), with higher scores indicating greater neglect severity. The maximum total score is 30, reflecting the overall impact of neglect on daily activities. According to Azouvi et al. (2002, 2003), the CBS score is used to classify neglect severity as follows: 0 = no behavioral neglect, 1–10 = mild behavioral neglect, 11–20 = moderate behavioral neglect, 21–30 = severe behavioral neglect. The two ADL tests NET and CBS were carried out at time points E1 and E4. In addition to the ADL tests, a further test for spatial neglect was applied at these time points. The FET is an exploration task performed on a Samsung S7+ tablet ^27^. During the familiarization phase, patients were required to locate a virtual origami bird in real space, which was augmented into the environment via the tablet’s rear camera. In the subsequent test phase, patients were asked to search for the bird again, even though it had not been hidden. The patients’ exploratory movements were recorded for 30 seconds, and the average horizontal exploration movement was measured. Values exceeding 9° indicate the presence of spatial neglect. The decision to conduct the FET at only two time points (E1 and E4) was due to time constraints and economic considerations within the rehabilitation setting, as therapy sessions were tightly scheduled and testing time was limited. The four traditional neglect tests (Letter Cancellation Test, Bells Test, Copying Task, Line Bisection Task) were prioritized for repeated administration at every assessment point to ensure comparability with previous studies that have widely used these standard assessments.

## Results

The potential confounding variables age, sex, and post stroke interval did not significantly differ between the active NMV group and the placebo NMV group (age: t(18)=-0.21, p=0.84; sex chi-square (1, N= 20)= 0.2, p=0.65; post-stroke interval t(18)=1.29, p=0.21), and therefore were not included in further analyses. Figure 3 provides an overview of the results for both the active NMV group and the placebo NMV group across all diagnostic examinations for the four diagnostic neglect tests. In both groups, no significant differences were found between the measurement time points E1 and E2 for any of the four neglect tests (active NMV group: dependent t tests for all four neglect tests: p>0.2; placebo NMV group: for all four neglect tests: p>0.4). This suggests that spontaneous remission can be ruled out as a factor when treatment started. For the subsequent analyses, we averaged the two measurements obtained at time points E1 and E2 into a single (“baseline”) variable, separately for each of the four diagnostic neglect tests.

**Figure 3.**
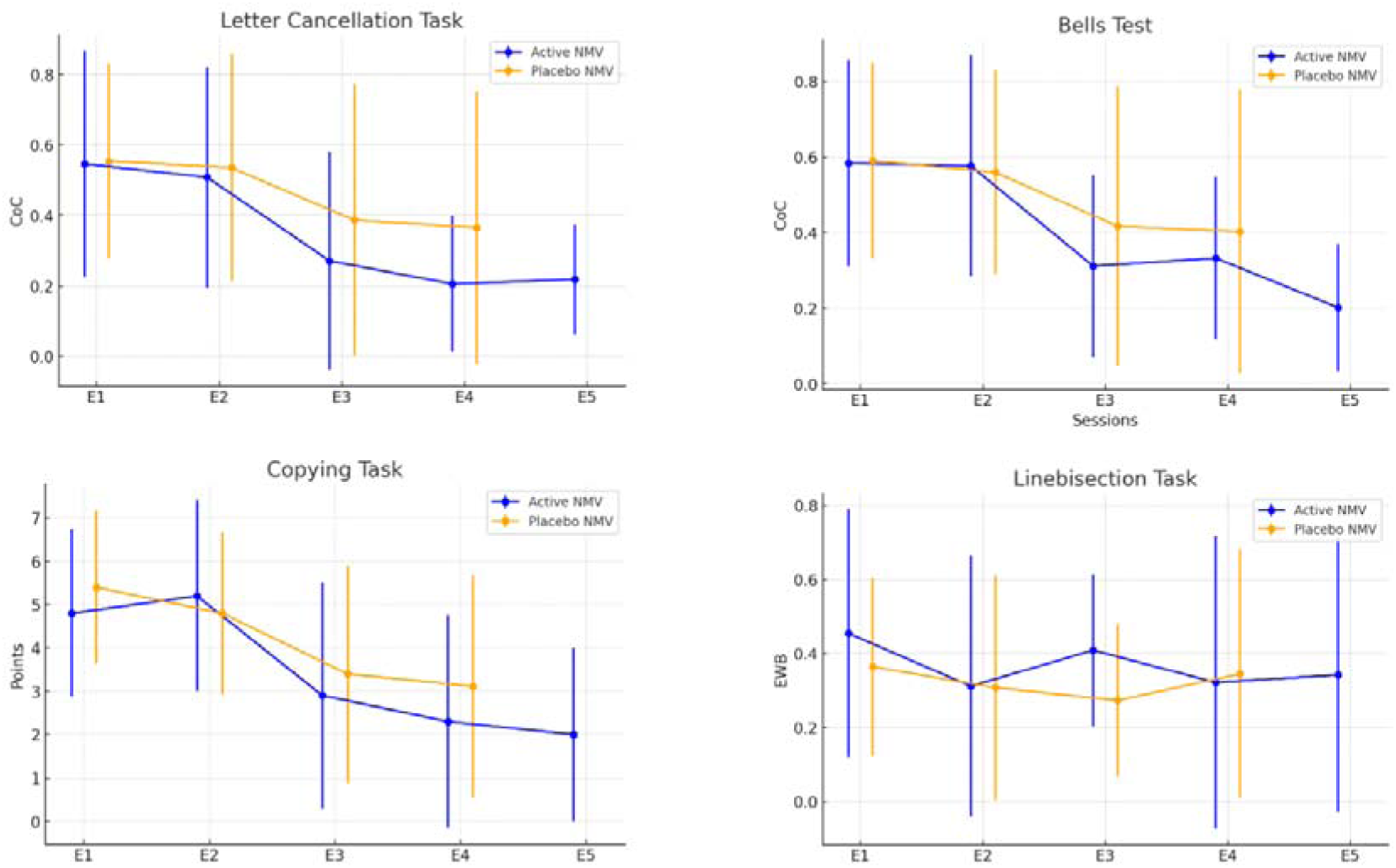
Results in the Letter Cancellation Task, Bells Test, Copying-Task, and Line bisection Task for the active NMV group (blue) and the placebo NMV group (orange) over the different diagnostic examination time points (E1 to E5).

### Treatment Effects

To assess treatment effects within each group, we conducted pairwise t-tests with a Bonferroni-corrected significance level of p<0.017 for pre- and post-intervention comparisons. For the four diagnostic neglect tests (Letter Cancellation Task, Bells Test, Copying Task, and Line Bisection Task), pairwise t-test compared “baseline” (average of E1 and E2) with E3. The results revealed significant improvements in three out of the four neglect tests in the active NMV group, whereas the placebo NMV group showed a significant change in one of the four neglect tests, namely the copying task (see Table 2). To investigate whether or not the experience of a visual illusion prior to stimulation influenced treatment outcomes, we also performed independent-samples t-tests on difference scores (“baseline” to E3) within the active NMV group for each of the four conventional neglect tests. Results revealed no significant differences (all p > 0.124).

**Table 2.** Treatment effects for the active NMV group (left) and the placebo NMV Group.

| Neglect Diagnostic Tests | Active NMV group |  |  |  | Placebo NMV group |  |  |  |
| --- | --- | --- | --- | --- | --- | --- | --- | --- |
|  | T | df | *p* | Cohens *d* | T | df | *p* | Cohens *d* |
| Letter Cancellation Test | 2.58 | 9 | .015* | 0.82 | 2.18 | 9 | .028 | 0.69 |
| Bells Test | 4.1 | 9 | .001* | 1.3 | 1.94 | 9 | .042 | 0.61 |
| Copying Task | 3.99 | 9 | .002* | 1.26 | 5.25 | 9 | <.001* | 1.7 |
| Linebisection Task | -.27 | 9 | .394 | -0.09 | 1.57 | 8 | .077 | 0.52 |
| Free-Exploration Test | 7.37 | 9 | <.001* | 2.33 | 2.422 | 8 | .021 | 0.81 |
| **Activities of Daily Living** |  |  |  |  |  |  |  |  |
| **NET** |  |  |  |  |  |  |  |  |
| Reading newspaper | -7.1 | 9 | <.001* | -2.25 | -3.2 | 9 | .005* | -1.01 |
| Writing an address | -3.15 | 9 | .006* | -1 | -2.57 | 9 | .015* | -.81 |
| Picture descriptions |  |  |  |  |  |  |  |  |
| *Menu* | -3.09 | 9 | .006* | -.98 | -1.58 | 9 | .075 | -.5 |
| *Sink* | -5.58 | 9 | <.001* | -1.76 | -3.24 | 9 | .005* | -1.03 |
| *Room* | -7.42 | 9 | <.001* | -2.35 | -3.18 | 9 | .006* | -1 |
| Reading a menu | -4.33 | 9 | <.001* | -1.37 | -4.21 | 9 | .001* | -1.33 |
| Reading the time |  |  |  |  |  |  |  |  |
| *Digital* | -.96 | 9 | .181 | -.3 | -2.9 | 9 | .009* | -.92 |
| *Analoge* | -2.74 | 9 | .011* | -.87 | -4.3 | 9 | <.001* | -1.36 |
| **Catherine Bergego Scale** | 3.23 | 9 | .005* | 1.02 | 3.52 | 9 | .003* | 1.11 |

(right).

In addition to the four diagnostic neglect tests, a significant improvement in exploratory behavior was also observed in the FET from E1 to E4 in the active NMV group; in contrast, the placebo NMV group showed no significant change in exploration behavior. Regarding the ADL test battery (NET), both groups exhibited significant improvements from E1 to E4 in 7 out of 8 tasks, as well as in the observer-rated CBS scale (see Table 2).

### Development over time

Pairwise comparisons within the active NMV group revealed no significance difference between E3 and E4 for any of the four neglect tests (all *p*>0.05) and no significant changes between E4 and the follow-up examination E5 (all *p*>0.09). This suggests that the improvements achieved through NMV remained stable over time. Similarly, the placebo NMV group with standard neglect treatment showed no significant difference between time points E3 and E4 for all five neglect tests (all *p*>0.13).

### Between-group effects

To examine whether the two groups differed from each other, a series of ANCOVAs were conducted in the first step, comparing “baseline” to E3 for the four neglect tests (Letter Cancellation Task, Bells Test, Copying Task, and Line Bisection Task) as well as the two time points before and after therapy for the FET and the ADL assessments (NET, CBS). For the NET, an averaged variable was computed across all eight tests. The ANCOVAs revealed no significant group differences for any of the four neglect tests (Letter Cancellation Task: F(1,17)=0.69, p = 0.42; Bells Test: F(1,17)=1.08, p=0.31; Copying Task: F(1,17)=0, p=0.99; and Line Bisection Task: F(1,16)=1.33, p=0.15). Similarly, there was no significant difference for the FET (F(1,16)=0.31, p=0.58) and the ADL assessments, including the averaged NET variable (F(1,17)=0.12, p=0.74) and CBS (F(1,17)=0.98, p=0.34). This indicates that both groups improved to a similar extent in terms of neglect severity, exploratory behavior, and everyday functioning, suggesting that the therapeutic effects were comparable across groups.

As a secondary analysis, a linear mixed model (LMM) was applied to the four neglect tests at each time point (E1–E4), as this approach effectively handles missing values. Missing values occurred only in a few individual cases due to specific circumstances. (In one case, the Line Bisection Task (E1–E4) was not completed because the patient was unable to perceive the line. Another patient was unavailable for testing at E1 for the FET due to an external appointment, which also prevented completion at E4. In a third case, all four neglect tests at E4 were not conducted because the patient experienced severe pain and opted for early termination.) The LMM analysis confirmed that the groups did not significantly differ at any of the test time points (E1–E4) for any of the four neglect tests (all p>0.29).

## Discussion

In a randomized, controlled, and blinded trial, this study investigating the isolated effects of neck muscle vibration (NMV) compared to standard neglect therapy. Both treatment approaches led to significant improvements of spatial neglect. While the active NMV group showed significant improvements in three out of four standard neglect tests (Letter Cancellation Task, Bells Test, Copying Task), the placebo NMV group exhibited improvement in only one of these tests (Copying Task). Additionally, both groups demonstrated substantial gains in exploration activity and ADL performance, as measured by the Free Exploration Test (FET), the Neglect-Test (NET), and the Catherine Bergego Scale (CBS). Direct comparisons between the two groups revealed no statistically significant differences, indicating that both NMV and standard neglect therapy were equally effective. Importantly, while both groups were re-assessed one week after therapy completion, the active NMV group underwent an additional follow-up one month post-treatment. The improvements in the active NMV group remained stable also over this longer time range.

### Methodological and Clinical Context

A recent review by Duclos et al. (2024) characterized the research landscape on NMV in neglect as methodologically inconsistent and acknowledged that several studies were of lower quality. A central limitation identified was the absence of randomized controlled trials that isolated the effects of NMV. The present study addresses this gap and proved that NMV is similarly effective as active exploration training, i.e., the treatment strategy currently considered the “gold standard” in neglect rehabilitation (cf. ^8,30^).

Moreover, the present study addresses another deficit identified by Duclos et al. (2024), namely the lack of ecologically valid outcome measures and the lacking of translational applicability. In fact, this aspect is of high clinical relevance, as persistent everyday functional limitations can remain even when core symptoms remit ^31^. By using functionally relevant instruments (CBS, FET, subtests from the NET), our study demonstrated that NMV indeed induces not only improvements on diagnostic tasks but also meaningful functional changes in patients’ exploration activity and ADL performance. This aligns with prior findings by Schindler et al. (2002), who applied NMV in patients with spatial neglect and found gains in reading performance and in a self-developed ADL observer questionnaire. Also, Kamada et al. (2011) reported improvements on the Japanese version of the Behavioral Inattention Test (BIT) which comprises 15 subtests - 6 conventional neglect tests (e.g., line bisection, cancellation tasks) and 9 assessing activities of daily living (ADL).

### Practical Relevance and Implementation Considerations

A major advantage of NMV is the passive application of the therapy from the patient’s point of view. Especially in the acute and subacute phases after stroke—when patients are often unable to engage in active exploration tasks or cognitive compensation strategies—such an intervention offers a valuable therapeutic approach. The fact that NMV requires no active participation of the patient and yet induces measurable functional improvements makes it an ideal building block especially (but not only) for early rehabilitation ^8^.

Importantly, our data suggest that the precise vibrator placement with the goal of inducing a visual illusion is (although desirable) not a prerequisite for clinical effectiveness. Of the ten patients in the active NMV group, five reported experiencing such a kinesthetic illusion of the centrally presented light stimulus. Nevertheless, significant improvements in exploratory behavior occurred even in the absence of consciously perceived illusions. We did not find significant differences between those participants who did vs did not report perception of a visual illusion while searching for the exact placement of the vibrator. This finding aligns with previous reports indicating that NMV despite successful manipulation of wanted behaviour produces a visual illusion of light spot displacement in only about 65 - 90% of subjects ^11,12,17,32–34^. With regard to this apparent inconsistency Seizova-Cajic & Sachtler (2007) argued that the computation of the egocentric spatial reference frame by NMV takes place at a higher level of the visual system and is not based on the conscious perception of motion or on eye movements. Rather, proprioceptive signals from the neck muscles appear to be directly fed into multimodal areas—particularly within the parieto-temporal cortex—where they contribute to the spatial localization of objects in reference to own body ^35^. Illusions resulting from this process can therefore remain unconscious, but still exert a functional influence ^36^.

The finding that the exact placement of the vibrator on the left posterior neck muscles is not a prerequisite for clinical efficacy, significantly lowers the practical barrier to entry for clinical use and increases the accessibility of NMV in routine care. It appears that accurate placement of the vibrator, as implemented in our patient sample, is sufficient for clinical efficacy: the vibrator was positioned within a palpable depression in the left dorsolateral neck region. Based on anatomical orientation, this site corresponds to the upper part of the so-called ‘posterior triangle of the neck’, i.e. the area of the depression between the left sternocleidomastoid muscle and the upper, descending part of the left trapezius muscle.

### New perspectives for muscle vibration in neglect: vibrating the biceps muscle

Interestingly, recent findings in healthy participants ^34^ have suggested that vibration of the right biceps muscle can induce a horizontal shift in SSA, similar in magnitude to that produced by left-sided NMV. This effect may stem from proprioceptive alterations in the limb-to-trunk signal ^34^. Such findings open up novel therapeutic possibilities for patients with spatial neglect: the right biceps muscle could potentially be targeted in future vibratory interventions. Also, simultaneously vibrating the right biceps and the left posterior neck muscles may turn out a promising approach to enhance therapeutic effects. Previous studies have demonstrated that stimulating multiple proprioceptive and sensory input channels simultaneously—such as neck muscle vibration, physical trunk rotation, and vestibular stimulation—can have additive effects. These combined stimulations, which play a role in shaping egocentric spatial coordinate systems, have been shown to amplify shifts in SSA and lead to greater reductions in neglect symptoms ^12,15,16,37^.

Also, the combination of bottom-up (e.g., left NMV, right biceps vibration) and top-down approaches (e.g., active exploration training) could possibly represent a suitable form for even better therapeutic success in neglect rehabilitation. Like Schindler et al. (2002) with using a combination of NMV with active exploration therapy, a recent single-case study by Ceyte et al. (2019) has suggested additive effects when NMV was combined with, e.g., arm movements.

### Conclusion and Outlook

The present study demonstrated that NMV, even when used in isolation, produces significant and lasting improvements in neglect symptoms and ADL-performance. The therapy is effective, functionally relevant, feasible, and comparable in efficacy to well-established approaches such as active exploration training. Moreover, it is a low-effort rehabilitation method that requires no active participation by the stroke patient and is therefore an ideal building block, especially (but not only) for early rehabilitation. In light of these findings, it seems imperative to integrate NMV more extensively into standard clinical practice and treatment guidelines.

Future research in NMV should focus on three key areas: (1) the (simultaneous) combination of NMV with top-down and/or other bottom-up therapy approaches (e.g., vibration of the biceps muscle), and (2) the development of portable, hands-free NMV devices as such systems could be worn continuously during other therapies.

## Data Availability

All data produced in the present study are available upon reasonable request to the authors

## Acknowledgements

We would like to thank L. Müller, O. Roggon and M. Wejt for their valuable contribution to data collection.

## Notes

### Competing Interest Statement

The authors have declared no competing interest.

### Clinical Trial

The study was preregistered in the WHO-approved public trial registry German Clinical Trials Register (DRKS-ID: DRKS00032376).

### Funding Statement

This study did not receive any funding

### Author Declarations

Ethics committee of University Hospital Tuebingen gave ethical approval for this work.

